# Antibody selection strategies and their impact in the analysis of malaria multi-sera data

**DOI:** 10.1101/2022.10.06.22280719

**Authors:** André Fonseca, Mikolaj Spytek, Przemyslaw Biecek, Clara Cordeiro, Nuno Sepúlveda

## Abstract

**Background:** Nowadays, the chance of discovering the best antibody candidates for explaining naturally acquired protection to malaria and detecting exposure to malaria parasites has notably increased due to publicly available multi-sera data. The analysis of these data is typically divided into a feature selection phase followed by a predictive one where several models are constructed for the outcome of interest. A key question in the analysis is to determine which and how each feature should be included in the predictive stage.

**Results:** To answer this question, we developed three approaches for classifying malaria protected and susceptible groups: (i) a basic and simple approach based on selecting antibodies via the nonparametric Mann-Whitney test; (ii) a dichotomization approach where each antibody was selected according to the optimal cut-off via maximization of the χ^2^ statistic for two-way tables; (iii) a hybrid parametric/non-parametric approach that integrates Box-Cox transformation followed by a t-test, together with the use of finite mixture models and the Mann-Whitney test as a last resort. We illustrated the application of these three approaches with published serological data for predicting clinical malaria in 121 Kenyan children. The predictive analysis was based on a Super-Learner where predictions from multiple classifiers were pooled together. Our results led to almost similar areas under the Receiver Operating Characteristic curves of 0.72 (95% CI = [0.61, 0.82]), 0.80 (95% CI = [0.71, 0.90]), 0.79 (95% CI = [0.7, 0.88]) for the simple, dichotomization and hybrid approaches, respectively.

**Conclusions:** The three feature selection strategies provided a better predictive performance of the outcome when compared to the previous results solely relying on Random Forests alone (AUC=0.68). Given the similar predictive performance, we recommended the three strategies should be used in conjunction in the same data set and selected according to their complexity.

## Background

Multi-sera data, where multiple antibody targets are measured in blood samples from the same individual, are becoming widely available in malaria research due to substantial developments at the level of serological assays^1–4^. This public availability has boosted basic research on the discovery of key antibodies associated with protection to malaria^5–10^. Moreover, it also motivated the development of serological-based algorithms that could predict not only past exposure to malaria parasites^11,12^ but also time since the last infection^13^. It has been suggested that these algorithms could help to design better malaria control strategies such as the serological testing and treatment (seroTAT) approach based on 8 antibodies for detecting Plasmodium vivax cases that should be targeted to receive an anti-hypnozoite therapy^12^.

In these multi-sera studies, the total number of antibody targets varied from dozens^8,10,13^ to thousands^6,7,14^. This number generates a huge computational cost for algorithms that search for the best model for the data. To overcome this problem, a brute-force approach (where every possible antibody combination is tried out) is computationally feasible for no more than 5 antibody targets^8^. However, it is not recommended above that number^10,12^.

This computational drawback motivated the use of data analysis strategies that are generically divided into an antibody or feature selection stage, followed by a predictive one, in which several statistical or machine learning models are estimated from the data^7,9,10,13,15^. In this scenario, the initial antibody selection stage determines the predictive performance of the models to be constructed in the following stage.

Antibody selection can be formulated as the procedure to determine which antibodies are associated with the outcome of interest^16–18^. However, this question hides another one in which one should decide whether data transformation, including dichotomization, should be used. Data transformation is particularly relevant in multiplex serological assays because distinct data distributions might emerge due to differences in the calibration curves across antibodies, as demonstrated in assay-optimization studies^16–18^. Until now, antibody selection has been carried out using only raw or untransformed^5,6^ data or seroprevalence-like data but^10,12^ without any combination of both. Additionally, the transformation of each antibody data is typically not considered. Therefore, current antibody selection procedures for multi-sera data lacks the flexibility to accommodate different data patterns. The current study tackles this issue and shows that it can potentially increase the chance of obtaining improved outcome predictions.

This paper presents a general parametric strategy for antibody selection in which a combination of transformed and dichotomized antibody data can be selected for the predictive phase. This strategy add flexibility to feature selection by combining the Box-Cox data transformation with well-known parametric statistical tests. A second, and simpler, strategy is also presented in which data of each antibody is initially dichotomized using an optimal cut-off point in the antibody distribution based on the maximization of the chi-square test statistic. Lastly, the predictive performance of these strategies was compared with the performance of the basic approach, where the statistical significance for the nonparametric Mann-Whitney-Wilcoxon test was obtained for each antibody, comparing the protected and susceptible groups. To illustrate these three strategies, we analyzed a published dataset on Immunoglobulin G (IgG) antibody responses to 36 Plasmodium *falciparum* (*Pf)* antigens in Kenyan children to understand protection to clinical malaria^8^ and whose data analysis was previously done with Random Forest^15^.

## Methods

### Data under analysis

We re-analyzed published data of 121 Kenyan children (age range: 1-10 years of age) described in detail elsewhere^8^. All children had a documented parasitaemia (parasite-positive) at the time of sampling and were monitored for clinical episodes of malaria over a follow-up period of 6 months. As in the original publication, children were considered susceptible (Sus, ns=40) or protected (Prt, np=81, 33%) if they had or did not have any clinical episode during the follow-up period. The serological data referred to individual IgG antibody responses to 36 Plasmodium falciparum antigens. These antibody responses were measured by multiple enzyme-linked immunosorbent assays (ELISA). Detailed information about recruitment, study design and experimental protocols, among other aspects of this early research, can be found in the original publication^8^.

### Preliminary antibody feature selection using Random Forest

The Random Forest works by constructing multiple decision trees trained on different parts of the same training set by a resampling process called bootstrap aggregation or bagging^19^. Random Forests were implemented by repeatedly fitting the model to 1000 resampled subsets of the data (100 repeats of 10-fold cross-validation). For each repetition, the dataset was divided into 10-cross-folds, of which 9-folds were used to perform an inner 10-fold cross-validation^20^. The number of trees to grow and the number of predictors randomly sampled as candidates in each split was set to default^21^ (number of trees = 500; number of predictors randomly selected = 2, 19 and 36), and the optimization criteria was maximization of the area under the of Receiver Operating Characteristic (ROC) curve, known as AUC^22^. Feature importance was given by the mean decrease in accuracy^23^. Briefly, for each tree, the prediction accuracy on the out-of-bag portion of the data is recorded. Then, after permuting each predictor variable, the prediction accuracy on the out-of-bag portion of the data is once again recorded. The difference between the two accuracies are then averaged over all trees, and normalized by the standard error^23^.

### Antibody selection based on the simple approach

The first antibody selection strategy was to select the antibodies by their statistical significance according to the non-parametric Mann-Whitney-Wilcoxon test comparing the protected and susceptible groups in each antibody data^24^.

### Antibody selection based on the data dichotomization approach

The second antibody selection strategy was based on a procedure in which the optimal cut-off to differentiate one study group from another was estimated by maximizing the chi-squared (χ^2^) statistic for testing independence in two-way contingency tables, as done elsewhere^25,26^ (Figure 1). In more detail, the values of each antibody were sorted by increasing order and then used to divide individuals into two latent serological groups (i.e., seronegative/seropositive individuals or high/low responders). For each value of a given antibody, the resulting data was summarized into a two-way contingency table comprising the qualitative variables serological status (below/above the cut-off) and malaria protection status (protected/non-protected). The χ^2^ test statistic was then calculated for this contingency table. After repeating this procedure for all antibody values, the optimal cut-off was selected as the value that maximized that test statistic, meaning the one that provided the best discriminatory ability between both groups of patients. After selecting the optimal cut-off, we calculated the respective *p-value* associated of the χ^2^ test. The dichotomized data was then used for the predictive phase. Finally, this procedure was repeated for each of the 36 antibodies included in the dataset.

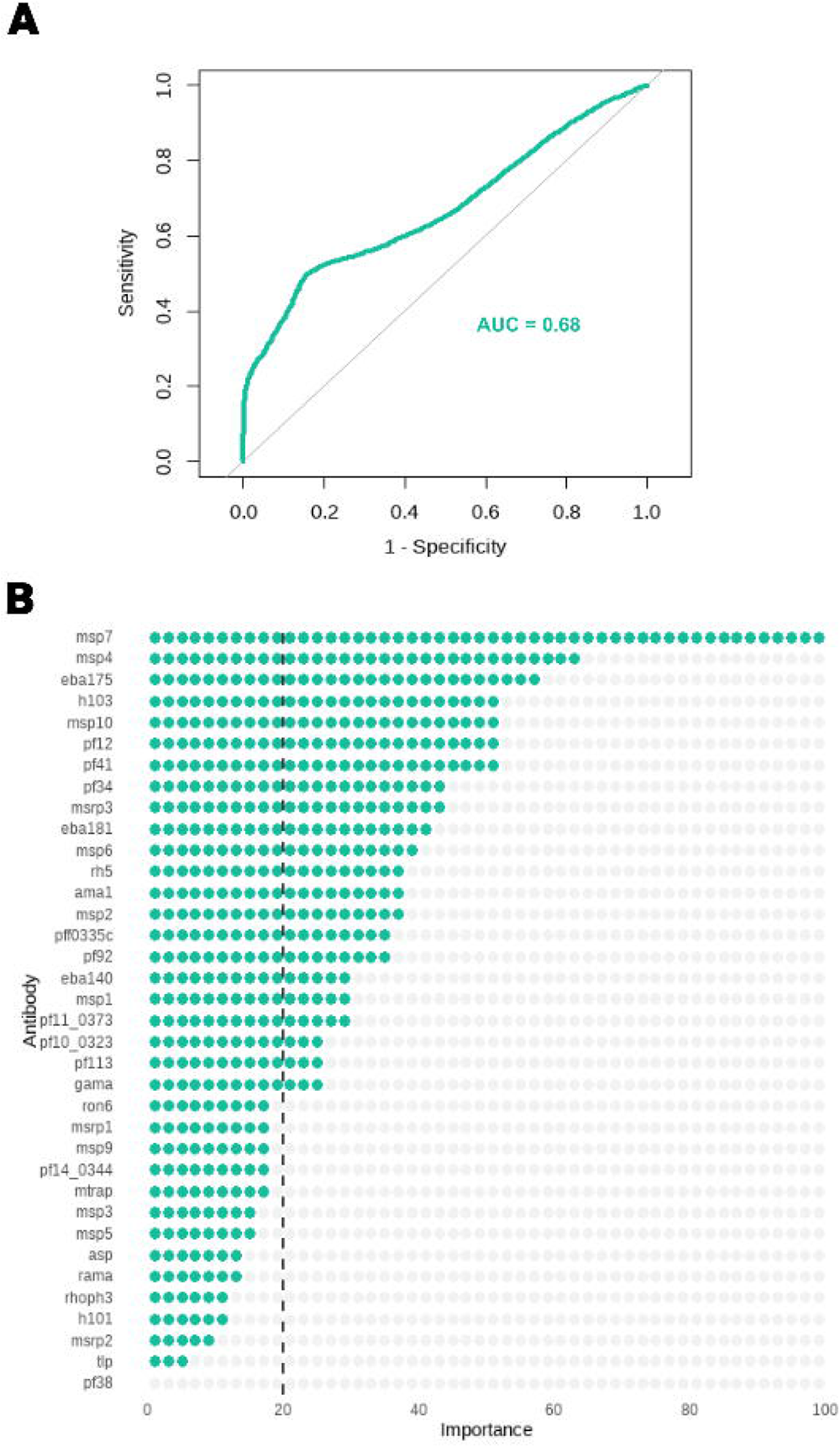

### Antibody selection based on the hybrid parametric/non-parametric approach

We adopted an alternative antibody selection approach using different parametric models or statistical tests (Figure 2). In the first step, we determined the optimal Box-Cox transformation for each antibody. This transformation was sought to obtain normal distributions with homogeneous variances in both groups. We searched the best parameter of this transformation (hereafter denoted as λ) within the interval (-4;4) by maximizing evidence for a Normal distribution using the Shapiro-Wilk (SW) test where the null hypothesis states that the data come from a normal distribution (with unknown parameters)^27^. A significance level of 5% was specified to assess whether the data of each antibody could follow a normal distribution.

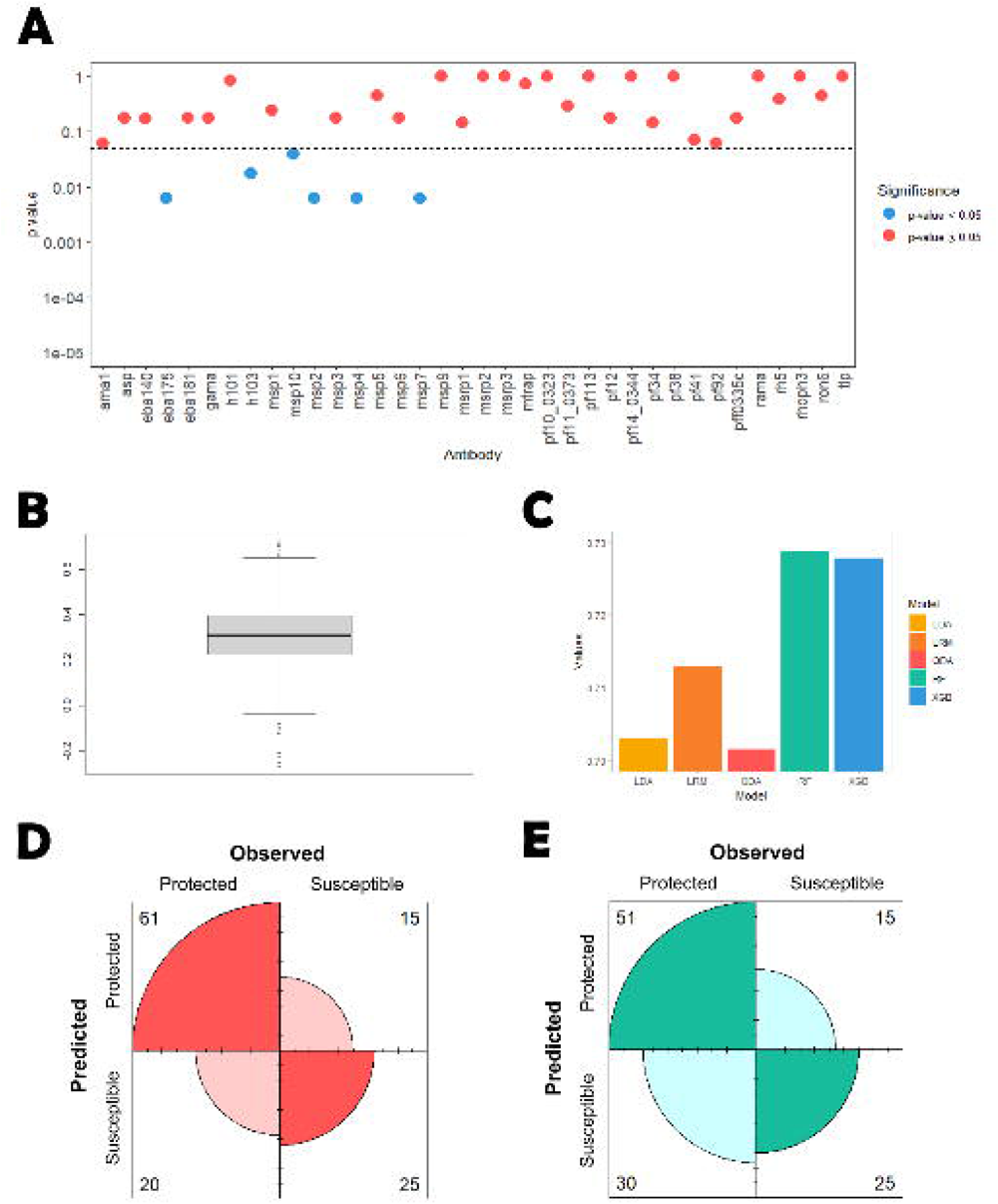

For the antibodies where there was no evidence against the normal distribution, we calculated the *p-value* for the t-test aiming at comparing the mean values of the susceptible and protected groups. The remaining antibodies, where there was evidence against the normal distribution, were then evaluated via finite mixture models given that it is recurrent to find latent populations in serological data^28^. Using transformed data, we estimated two-component mixture models based on Normal, Generalized t, Skew-Normal and Skew-t distributions by maximizing the likelihood function via the Expectation-Maximization algorithm^29^. We also estimated the Generalized t, Skew-Normal and Skew-t distributions to assess the evidence that the data could come from a single non-Normal serological population beyond the ones identified by the Box-Cox transformation. We compared all these models using the Akaike’s Information Criterion (AIC) and performed the Pearson’s goodness-of-fit test by dividing the respective data into deciles (i.e., 10%-quantiles). Minimization of the AIC, together with a good fit to the data, at the significance level of 5%, was the criterion for selecting the best model. For antibodies whose data provided evidence of two latent serological populations, we divided the individuals into two latent serological groups using the optimal cut-off by maximization of the χ^2^ statistic (as described in the previous section). For the antibodies in which there was evidence of a single latent serological population antibody, we constructed two linear regression models using the antibody values as the response variable. The first model comprised only the intercept (i.e., not including any covariate), while the second model comprised the malaria protection status as the single covariate. We then computed the *p-value* of the Wilks’s likelihood ratio test to compare the two models at the significance level of 5%. The rejection of the null hypothesis suggested statistically significant differences between the two models being compared. Finally, antibodies that could not be fitted by any of the above parametric models were analyzed through a non-parametric Mann-Wilcoxon test to compare the median values of the protected and susceptible groups.

### Correction for multiple testing

In each antibody selection strategy, all the *p-values* obtained were adjusted to ensure a global false discovery rate (FDR) of 5%. This *p-value* adjustment was made via the Benjamini-Yekutieli procedure under a general dependence assumption between tests^30^. All antibodies whose adjusted *p-values* < 0.05 were carried to the predictive analysis.

### Predictive Stage

When analyzing data resulting from each antibody selection strategy, we adopted a Super-Learner (SL) approach to predict the malaria protection status of each individual^31,32^. In general, this approach aims to estimate different (mostly data-driven) classifiers whose individual predictions for each individual are combined into a pooled estimate via a weighted average calculated by cross-validation. To construct this pooled estimator, we used the following 5 classifiers for each set of antibodies selected: logistic regression model (LRM) with main effects only, Random Forest (RF), linear discriminant analysis (LDA), quadratic discriminant analysis (QDA), and extreme gradient boosting (XGB). For the antibodies selected by optimal dichotomization antibody selection strategy, we did not include linear and quadratic discriminant analyses in the SL algorithm because these classifiers are more appropriate for data containing quantitative predictors only.

To assess the quality of the final predictions, we estimated the ROC curve and its area (AUC)^22,33^. In addition, we calculated the confusion matrices where the rows and columns represented the predicted and the observed status of the individuals, respectively^34^. These confusion matrices were calculated using the point in the ROC curve that minimizes the distance to the point (0,1) related to the perfect classification of the individuals, here called ROC01 criterion^35^. From the standpoint of constructing a fair classifier^36,37^, we also determined the predictive performance by the point in the ROC curve in which sensitivity (protected) and specificity (susceptible) were equal approximately^35^. This criterion is hereafter denoted as SpEqualSe criterion^35^.

### Statistical Software

All statistical analyses were implemented in the R^38^ version 4.3.0 using the following packages: “AID” to perform Box-cox transformation and to compute the respective Normality tests^39^; “caret” to construct the confusion matrices^23^; “doParallel” for parallel processing and faster run times^40^; “dplyr” to better manipulate the data^41^; “ggplot2” to plot the data^42^; “ggrepel to avoid overlaid text on plots^43^; “lmtest” to perform the likelihood ratio test^44^; “MASS” for general analysis^45^; “mixsnsm” to estimate mixture models based on Skew-Normal and Skew-t distributions^46^; OptimalCutpoints to obtain the point in the ROC curve that minimizes of the distance to the point (0,1)^35^; “pROC” to estimate ROC curves^47^; “sn” to perform linear regression models based on Skew-Normal or Skew-t distributions for the residuals^48^; “SuperLeaner” to perform all the predictive analysis^31^; “tydir” to facilitate data manipulation^49^.

## RESULTS

### Preliminary analysis based on the Random Forest approach

Initially, a Random Forest model was implemented on all 36 antibodies replicating the approach conducted by Valleta and Recker^15^. According to our findings, we were able to reproduce the previously reported AUC of 0.68^15^ (Figure 3A). Looking at the feature importance values, we concluded that all except one of the 36 antibodies were required to achieve this predictive performance (Figure 3B). Nevertheless, a more thorough analysis of the feature’s importance values reveals that several features had very low importance values (below 20% importance) (Figure 3B). This led us to hypothesize that removing these features could improve the model’s performance. Therefore, three distinct *filter* strategies for feature selection were used.

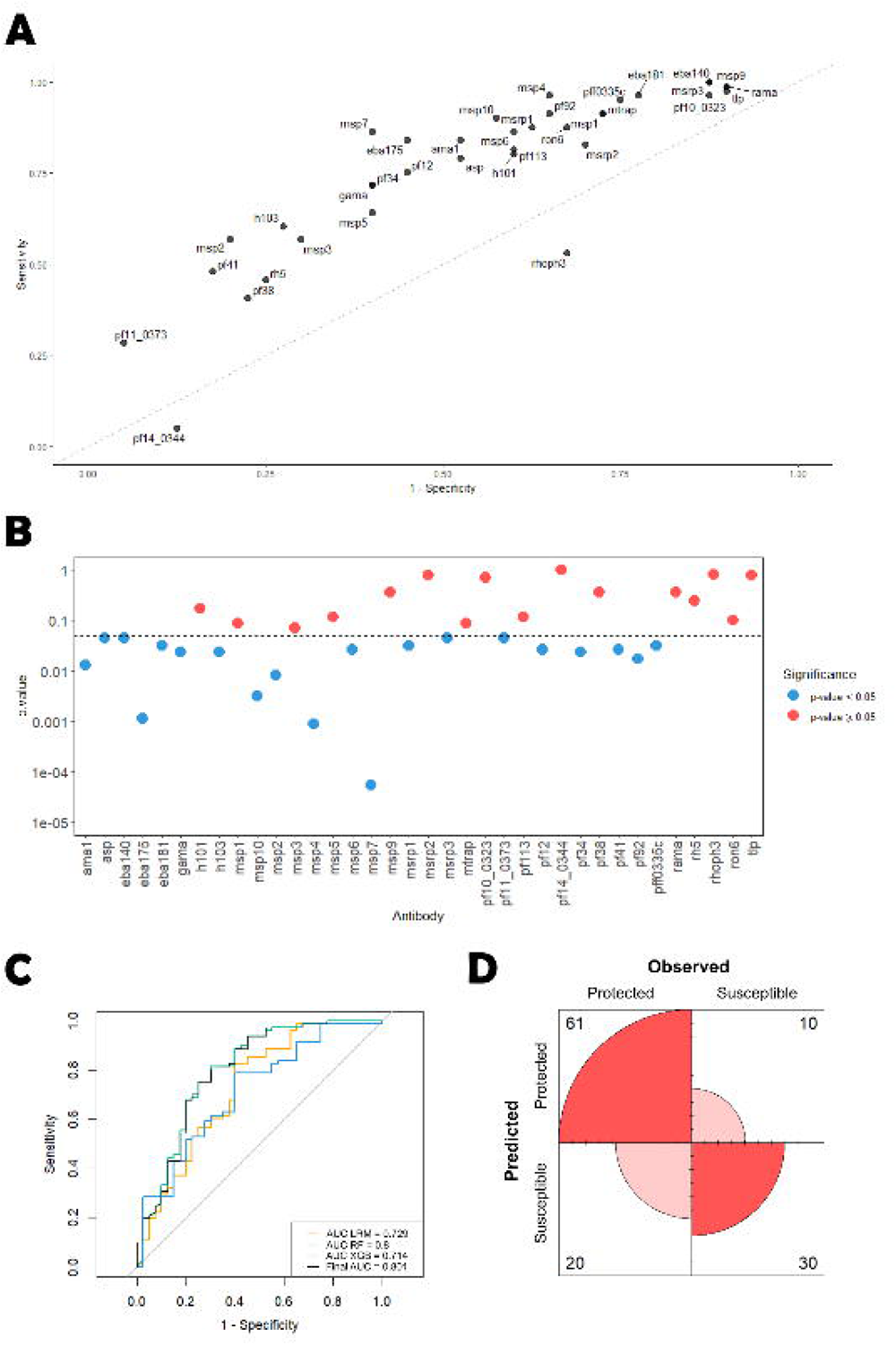

### Analysis based on the simple antibody selection approach

We first tested whether levels of each antibody were significantly different between susceptible and protected individuals using the Mann-Whitney-Wilcoxon test. According to this nonparametric test, twenty-one out of the 36 antibodies were found statistically significant before adjusting for multiple testing. This number dropped to six after controlling for an FDR of 5%: *msp2*, *msp4*, *msp10*, *eba175*, *msp7*, and *h103*. This substantial reduction in the number of significant antibodies is likely to be explained by the positive correlation among different antibodies (average Spearman’s correlation coefficient = 0.312; Figure 4A). As expected, the levels of these selected antibodies were elevated in the protected individuals (Figure 4B).

We then constructed a Super-Learner classifier based on the data of these 6 antibodies. The average estimates for the AUC were 0.713, 0.703, 0.702, 0.729 and 0.728 using LRM, LDA, QDA, RF and XGB, respectively (Figure 4C). The average weights of these classifiers were 0.089, 0.506, 0.035, <0.001, and 0.370 in the final predictions, respectively. These weights implied an AUC of 0.719 (95% CI=(0.615, 0.824)) for the SL predictions.

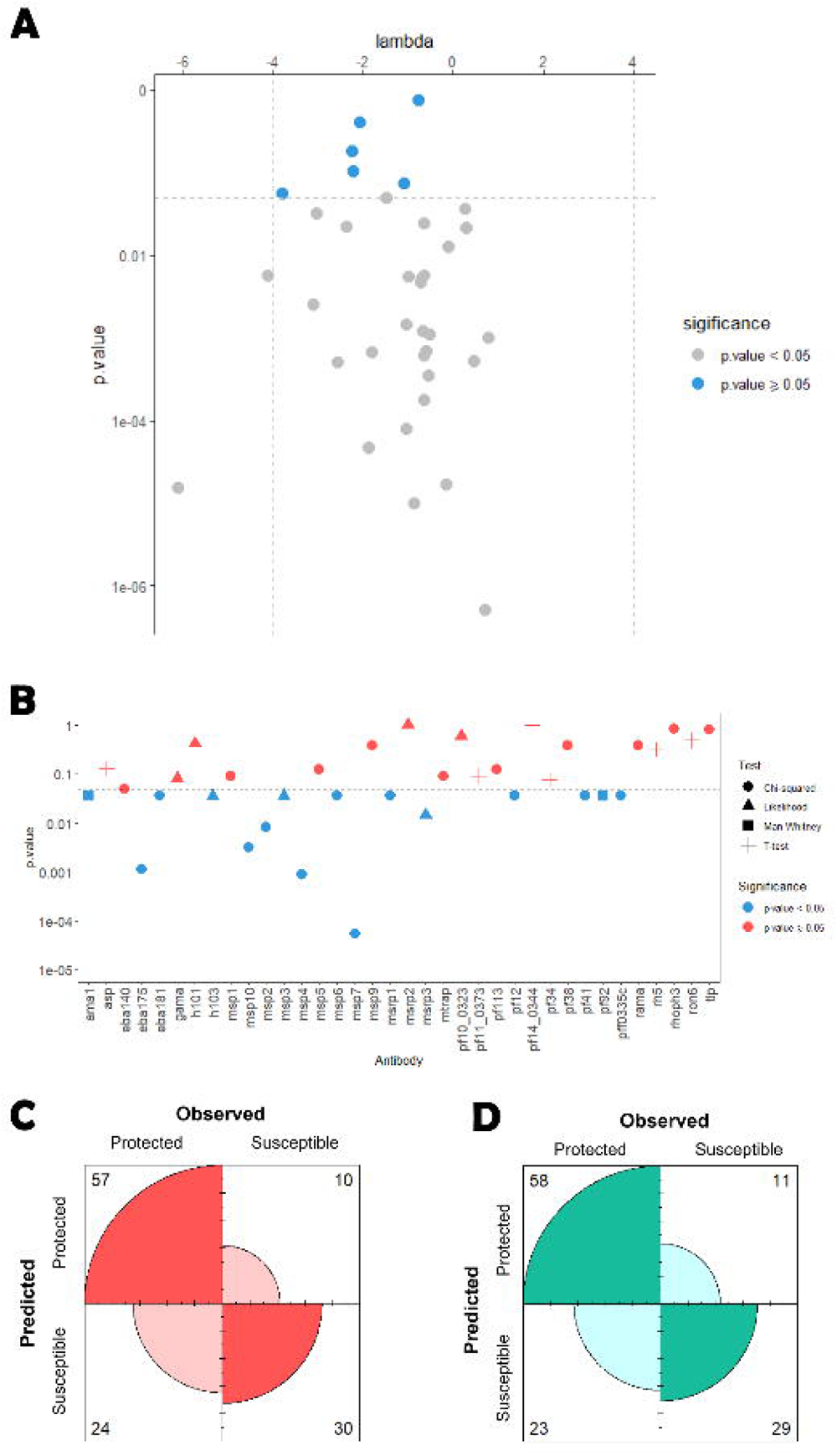

Moreover, the SL predictions had a sensitivity of 0.753 and a specificity of 0.625 according to the ROC01 criterion (Figure 4D). A higher number of protected individuals in the dataset could explain the fact that sensitivity was estimated at a higher value than specificity. To assess the final classifier without this potential selection bias, we determined the point at which the ROC sensitivity and specificity were similar and used it to obtain a fair classification (SpEqualSe criterion). The balanced sensitivity and specificity estimates were 0.630 and 0.625, respectively (Figure 4E).

### Analysis based the data dichotomizations approach

In this analysis, we determined the optimal classification cut-off for each antibody according to the χ^2^ statistics. The sensitivity using these optimal cut-offs varied from 0.049 (pf14_0344) to 1 (eba140, msrp3), while the specificity varied from 0.100 (msp9) to 0.95 (pf11_0373). The top 3 antibodies whose optimal cut-offs provided the sensitivity and specificity estimates closest to perfect classification (i.e., specificity=sensitivity=1) were msp7 (Se=0.852, Sp=0.600), eba175 (Se=0.827, Sp=0.550), and msp2 (Se= 0.556, Sp=0.800; Figure 5A).

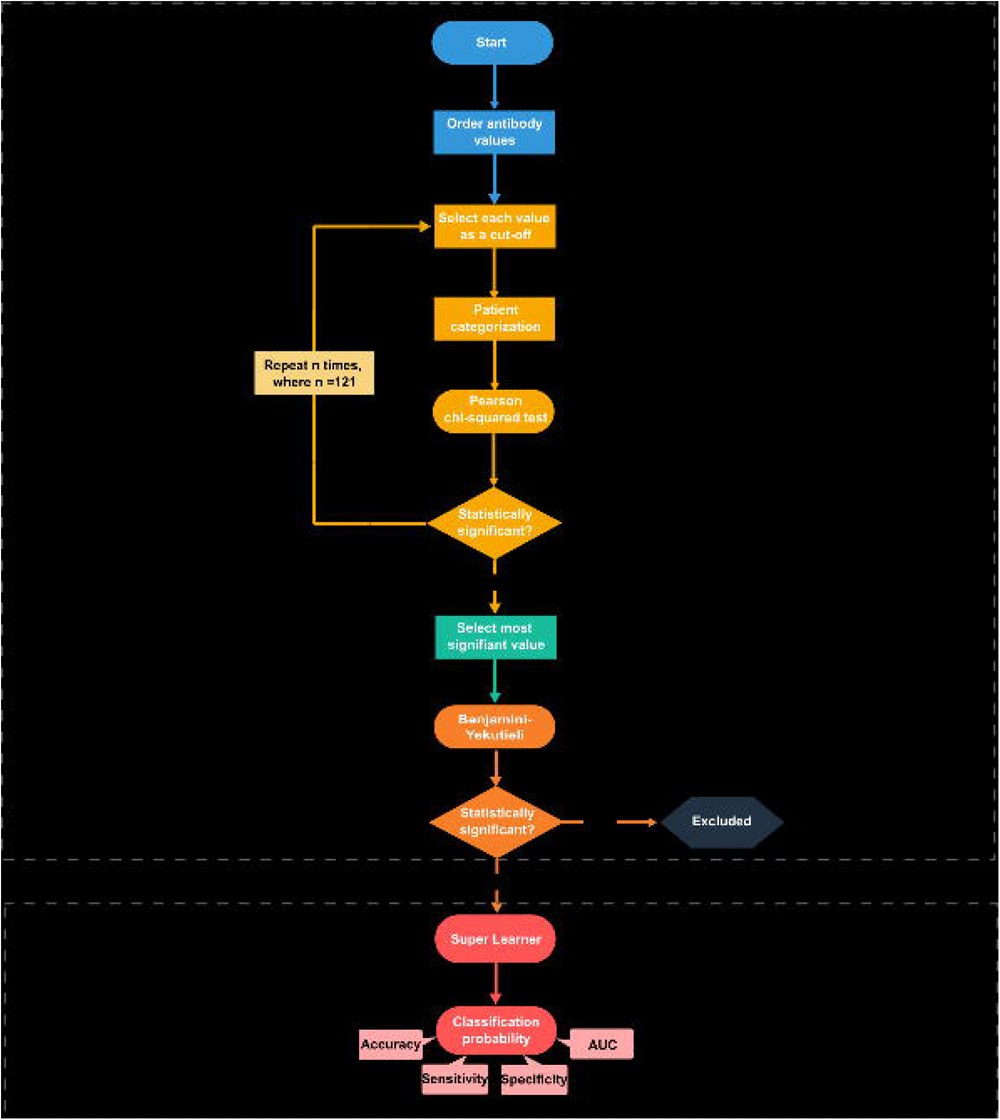

There were 28 out of 36 antibodies whose proportions above the optimal cut-off were significantly different between protected and susceptible individuals at the 5% significance level (Table 1). However, after controlling for an FDR of 5%, the number of statistically significant antibodies dropped to 20 (Figure 5B). The dichotomized data of these antibodies, according to the optimal cut-off, was used for predictive analysis.

**Table 1.** Patient Seroprevalence. The statistically significant results of the χ^2^ test for the 28 antibodies. The antibody levels that maximized the separation between the susceptible and protected group of individuals (Cut-off) and the proportion of seropositive individuals for all (Total), Protected (Prt) and susceptible (Sus) children, respectively

| Antibody | P-value | Cut-off | Total | Prt | Sus |
| --- | --- | --- | --- | --- | --- |
| msp1 | 0.01 | 0.15 | 0.85 | 0.91 | 0.73 |
| msp2 | <0.01 | 0.07 | 0.45 | 0.57 | 0.20 |
| msp4 | <0.01 | 0.13 | 0.86 | 0.96 | 0.65 |
| msp5 | 0.02 | 0.09 | 0.56 | 0.64 | 0.40 |
| msp10 | <0.01 | 0.25 | 0.79 | 0.90 | 0.58 |
| pf12 | <0.01 | 0.10 | 0.65 | 0.75 | 0.45 |
| pf92 | <0.01 | 0.11 | 0.83 | 0.91 | 0.65 |
| pf34 | <0.01 | 0.07 | 0.61 | 0.72 | 0.40 |
| pf113 | 0.02 | 0.05 | 0.74 | 0.81 | 0.60 |
| gama | <0.01 | 0.05 | 0.61 | 0.72 | 0.40 |
| ama1 | <0.01 | 0.16 | 0.74 | 0.84 | 0.53 |
| eba175 | <0.01 | 0.14 | 0.71 | 0.84 | 0.45 |
| eba140 | <0.01 | 0.11 | 0.96 | 1.00 | 0.88 |
| eba181 | <0.01 | 0.11 | 0.90 | 0.96 | 0.78 |
| mtrap | 0.01 | 0.05 | 0.85 | 0.91 | 0.73 |
| asp | <0.01 | 0.08 | 0.70 | 0.79 | 0.53 |
| msp3 | 0.01 | 0.08 | 0.48 | 0.57 | 0.30 |
| msp6 | <0.01 | 0.12 | 0.78 | 0.86 | 0.60 |
| msp7 | <0.01 | 0.24 | 0.71 | 0.86 | 0.40 |
| msrp1 | <0.01 | 0.05 | 0.79 | 0.88 | 0.63 |
| msrp3 | <0.01 | 0.04 | 0.96 | 1.00 | 0.88 |
| h101 | 0.03 | 0.05 | 0.74 | 0.80 | 0.60 |
| h103 | <0.01 | 0.07 | 0.50 | 0.60 | 0.28 |
| pf41 | <0.01 | 0.12 | 0.38 | 0.48 | 0.18 |
| pff0335c | <0.01 | 0.05 | 0.88 | 0.95 | 0.75 |
| rh5 | 0.04 | 0.16 | 0.39 | 0.46 | 0.25 |
| ron6 | 0.02 | 0.04 | 0.81 | 0.88 | 0.68 |
| pf11_0373 | <0.01 | 0.08 | 0.21 | 0.28 | 0.05 |

The AUC of the SL-based predictions was estimated at 0.801 (95% CI=(0.709, 0.892)) (Figure 5C), which showed an improvement from the previous analysis using a non-parametric antibody selection. The average AUC (and weights) estimates for each classifier were: LRM -0.729 (<0.001), RF -0.800 (0.973), and XGB – 0.714 (0.026). This result showed that, notwithstanding the reasonable AUC estimates for LRM and XGB, the final predictions were basically derived from the RF classifier. Note that LDA and QDA were not included in the SL algorithm, as they are more suitable for analyzing quantitative multivariate data.

According to the ROC01, the final sensitivity and specificity were estimated at 0.753 and 0.750, respectively. These estimates were identical for the SpEqualSe criterion. In conclusion, this analysis produced a combined classifier that exhibited an improved and better-balanced predictive performance than the previous one. However, this classifier had the disadvantage of including a higher number of antibodies comparing compared to the previous one (20 antibodies versus 6 antibodies).

### Analysis based on the hybrid parametric/non-parametric approach

We first estimated the Box-Cox optimal data transformation and applied it to the antibody data. Then, we compared the protected and susceptible groups using the parametric t-tests for two independent samples. Our findings suggested that there were 6 antibodies whose data in each study group could be analyzed by these tests after the Box-Cox transformation: *asp*, *pf11_0373*, *pf14_0344*, *pf34*, *rh5,* and *ron6* (Figure 6A); note that, at this stage, we did not adjust the *p-values* of the respective goodness-of-fit tests due to multiple testing, because such adjustment would increase the evidence for the null hypothesis of these tests. In these antibodies, the estimates for the parameter l of the Box-Cox transformation varied from -3.8 (*ron6*) to -0.78 *(pf34)*. The estimates suggest that the logarithmic transformation would not be the best to generate a normal distribution. The best evidence for a Normal distribution was found for *pf34* with a *p-value* of 0.75 using the Shapiro-Wilk test (Figure 6A).

The remaining 30 antibody data were then analyzed by fitting finite mixture models based on Normal, Generalized T, Skew-normal, and Skew-T distributions; note that Normal and t distributions come as special cases of the latter probability distributions. For the statistical convenience of having these antibodies defined in terms of positive and negative values, we log-transformed the respective antibody data.

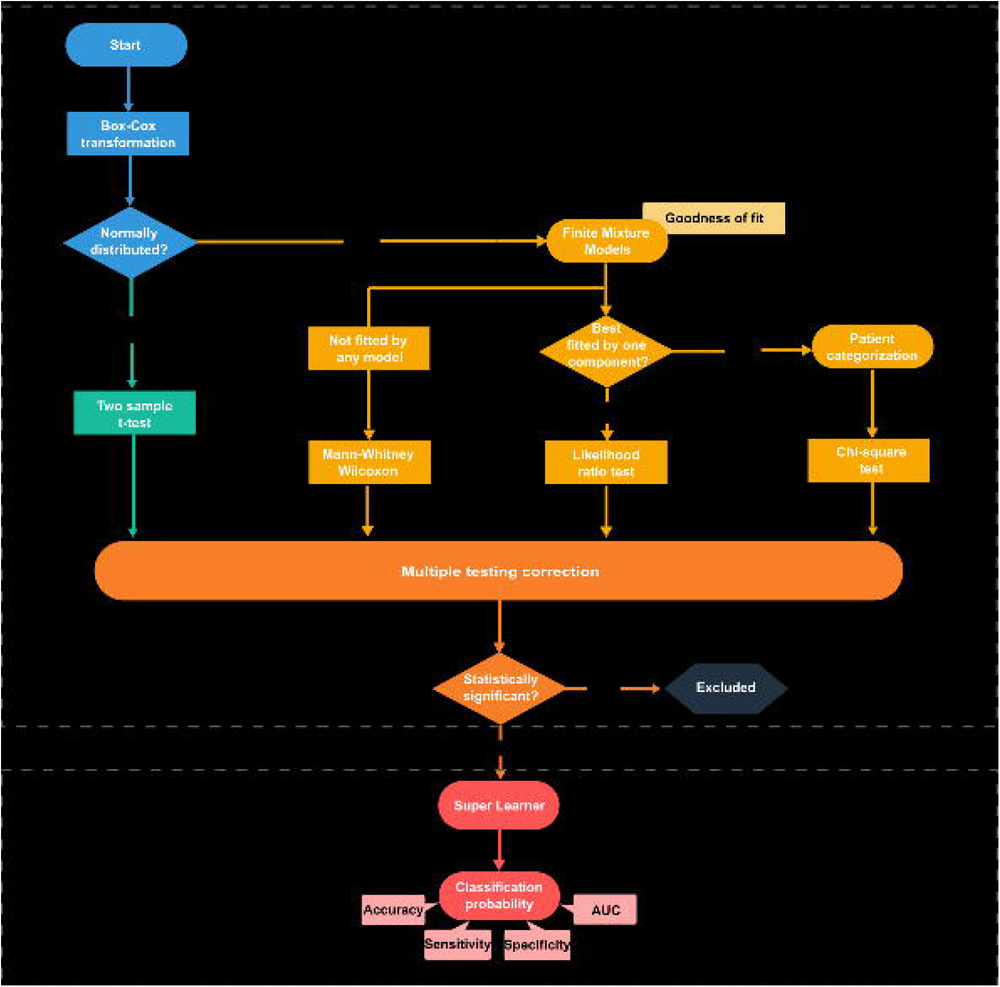

We found evidence that data from 7 antibodies could be described well by either Skew-Normal (*msp3* and *h103*) or Skew-t (*gama, h101*, *msrp2*, *msrp3*, and *pf10_0323*) distributions (Table 2). In this case, the comparison between study groups was made via regression models using these distributions for the errors. Except for the antibodies against *pf92* and *ama1,* data of the remaining antibodies were best described by a mixture of two Normal distributions (4 antibodies), two Skew-Normal distributions (16 antibodies) or two Skew-t distributions (1 antibody; see Table 2). The best fit of these mixture models was obtained for the antibody against *pf113* using a two-component Normal mixture model (p=0.73, Pearson’s goodness-of-fit test; Table 2). For these antibodies, we assumed the existence of a seronegative and a seropositive population. We dichotomized the respective data using the optimal cut-off by maximization of the χ^2^ test statistic. Data of the antibodies against *pf92* and *ama1* could not be fitted by either Normal distribution after Box-Cox transformation or using the above mixture models. Therefore, we used the Mann-Whitney-Wilcoxon test as the last resort statistical test to compare the protected and susceptible groups. Thus, comparing the protected and susceptible groups using the different tests led to 25 significant antibodies before applying a multiple testing correction This number decreased to 16 after ensuring an FDR of 5%. These antibodies were found to be significant by the Wilks’ likelihood ratio test (*msp3*, *msrp3* and *h103*), the c^2^ test (*eba175*, *eba181*, *msp2*, *msp4*, *msp6*, *msp7*, *msp10*, *msrp1*, pf12, *pf41*, *pff0335c*) and the Mann-Whitney-Wilcoxon test (*pf92, ama1*) (Figure 6B). In the predictive analysis, data of each antibody were included in the SL approach according to the suggested scale by the antibody selection procedure: log-transformed data for antibodies coming from the Wilks’ likelihood ratio test, dichotomized seropositive/seronegative data for antibodies coming from the c^2^ test, and the original scale for the *pf92* and *ama1*-related antibodies coming from the Mann-Whitney-Wilcoxon test. Before obtaining the combined predictions, we checked each individual classifier’s performance. The average AUC were 0.756, 0.807, 0.768, 0.656 and 0.643 using LRM, RF, LDA, QDA, and XGB, respectively. Therefore, the best individual classifier was the RF. The average weights of these classifiers were 0.021, 0.912, 0.0132, 0.053, and 0 in the final predictions, respectively, resulting in an AUC of 0.78 CI=(0.7, 0.879)) According to the ROC01 criterion, the sensitivity and specificity were estimated at 0.703 and 0.750, respectively (Figure 6C). Moreover, based on the ROC curve, the best balance between these quantities was obtained for a sensitivity and a specificity of 0.716 and 0.725, respectively (Figure 6D).

**Table 2.** Analysis based on Finite Mixture Model. Results from the analysis of 28 antibodies based on finite mixture models where AIC and GOF denote the Akaike’s information criterion and Pearson’s goodness-of-fit test, respectively.

| Antibody | Best Mixture Model | # Components | AIC | P-value (GOF) |
| --- | --- | --- | --- | --- |
| eba140 | Skew Normal | 2 | 23,92 | 0,32 |
| eba175 | Skew Normal | 2 | 33,29 | 0,03 |
| eba181 | Skew Normal | 2 | 42,9 | 0,03 |
| gama | Skew-t | 1 | -272,19 | 0,24 |
| h101 | Skew-t | 1 | -230,91 | 0,33 |
| h103 | Skew Normal | 1 | -41,91 | 0,72 |
| msp1 | Skew Normal | 2 | 25,35 | 0,26 |
| msp10 | Normal | 2 | 71,52 | 0,07 |
| msp2 | Skew Normal | 2 | -24,09 | 0,43 |
| msp3 | Skew Normal | 1 | 1,46 | 0,32 |
| msp4 | Skew Normal | 2 | 76,23 | 0,04 |
| msp5 | Normal | 2 | -71,25 | 0,33 |
| msp6 | Normal | 2 | -168,02 | 0,35 |
| msp7 | Skew Normal | 2 | 46,11 | 0,16 |
| msp9 | Skew Normal | 2 | -10,75 | 0,53 |
| msrp1 | Skew-t | 2 | -89,1 | 0,06 |
| msrp2 | Skew-t | 1 | -122,32 | 0,12 |
| msrp3 | Skew-t | 1 | -283,83 | 0,02 |
| mtrap | Skew Normal | 2 | -213,58 | 0,13 |
| pf10_0323 | Skew-t | 1 | -344,51 | 0,62 |
| pf113 | Normal | 2 | -139,5 | 0,73 |
| pf12 | Skew Normal | 2 | -33,29 | 0,24 |
| pf38 | Skew Normal | 2 | 99,41 | 0,05 |
| pf41 | Skew Normal | 2 | 35,96 | 0,10 |
| pff0335c | Skew Normal | 2 | 4,83 | 0,04 |
| rama | Skew Normal | 2 | -153,54 | 0,32 |
| rhoph3 | Skew Normal | 2 | -152,73 | 0,02 |
| tlp | Skew Normal | 2 | -426,93 | 0,02 |

## Discussion

Multi-sera data, where thousands of antibody targets are simultaneously measured, can increase the chance of discovering the best antibody candidates for explaining natural protection to malaria and detecting exposure to malaria parasites^50–52^. Nonetheless, this type of data brings novel challenges^53,54^. One of the main drawbacks when dealing with this type of data is the difficulty of identifying the relevant features for the task at hand. Among the thousands of features screened, most will be irrelevant or redundant and will negatively impact the predictive ability of a predictive model^55^. Not only that, trying to fit a predictive model to these many features increases the computational complexity and cost, reduces the model generalization ability, and affects the explainability of the model^54^. To overcome these limitations, feature selection strategies have been proposed, where the aim is to identify and remove all the irrelevant features so that the learning algorithm focuses only on those features of the training data useful for prediction^53^. This leads not only to a simpler interpretability, as when a small number of features is selected their biological relationship with the target disease is more easily identified, but also into a lower computational cost and increased accuracy stemmed from reducing the chance of overfitting^54,56^. Therefore, feature selection before the implementation of a predictive model is strongly advocated^57^. Amongst the different feature selection approaches, here we opted for the use of *filter* methods^53,56,57^. These rely on statistical measures (e.g *p-value,* correlation coefficient), and their application precedes the predictive phase, thus being independent of any predictive model^56,57^. For this reason, they are usually very fast to implement. Here we will discuss the advantages and drawbacks of the distinct filter methods employed in each proposed methodology. The simple approach relying on the Mann-Whitney Wilcoxon test for feature selection is the most scalable approach for larger datasets among the ones here proposed. It is the most straightforward and fastest approach to implement, making it an appealing tool for those looking for a low complexity model when conducting a classification task. Moreover, given its ranking intrinsic nature, this strategy represents the best option to achieve reproducible results^24^. Nevertheless, its low statistical and computational complexity comes at a cost since this feature selection approach might led to a lower predictive performance when compared to the other strategies, as demonstrated in this study. Concerning the data dichotomization methodology performance, we can see that, unsurprisingly, it was the one with better predictive performance and therefore, this methodology can be seen as the optimistic case scenario. The use of the χ^2^ test for feature selection is very straightforward to implement and has a low computational complexity making it an extremely attractive option for feature selection. However, this approach is highly dependent on the calculation of the optimal cut-off value for each antibody. Therefore, small perturbations in the data might influence its calculation, which reduces the chance of obtaining the same antibody selection and classifier. Thus, reproducibility may be hindered when using this feature selection methodology across different studies. Finally, despite being statistically challenging, our parametric approach provides a more comprehensive analysis of the data. In this approach, feature selection is made on the basis of data transformation and dichotomization via mixture modelling, thus accommodating different data patterns. However, this feature selection strategy is expected to increase the computational time dramatically with as the number of antibodies under analysis increases. The computational implementation in user-friendly packages is also not trivial in relation to the other feature strategies applied in this study. Finally, this feature selection strategy is based on complex statistical models such as finite mixture models related to Skew-Normal distributions. In this scenario, this strategy seems less appealing to the malaria research community where, despite the efforts to improve mathematical modelling capacity, the availability of qualified staff with statistical and machine learning skills remains scarce. Therefore, the use of simple filter methods seems a more viable solution at the moment, especially, when it comes to analyze data featuring thousands of antibodies. Such a case is seen in Proietti et al.^7^ where antibodies with a p-value < 0.01 for the univariate logistic regression were selected after Bonferroni correction followed by sparse partial least squares discriminant analysis (sPLS-DA) and Support Vector Machine (SVM). Another example is the use of the Spearman’s correlation coefficient to remove highly correlated antibodies prior to implementation of the Random Forest presented by Valletta and Recker^15^.

A significant disadvantage of *filter* methods is the inability to detect complex relations between multiple features and the outcome of interest, which generally translates into pourer results in the predictive phase^56,57^. Thus *wrappers* or *embedded methods* are more appealing. *Wrappers* are created around a particular classifier and rely on the classifier’s information concerning feature relevance^56,57^. For this reason, the computational effort they require is usually significant, becoming unfeasible in real time when thousands of features are considered. For this reason, *wrappers* are often avoided, and their implementation for feature selection in malaria is scarce^8^. A more attractive approach are embedded methods that use the core of a classifier to establish a criterion to rank features^53,56^. Embedded algorithms perform feature selection during the classifier training procedure while optimizing the feature set used to achieve the best accuracy. Therefore, they are less computationally costly than wrappers while still dealing with the complex interactions between multiple features and the outcome^53,56^. Examples of *embedded* feature selection methods intending to unveil antibody immune signatures in malaria are described in the literature. Aitken et al.^58^ used an elastic net-regularized logistic regression for antibody selection followed by a partial least squares discriminant analysis to find a minimal set of antibodies that accurately classified the individuals under analysis. Helb et al.^13^ used a hierarchical criterion for feature selection, where a combination of *embedded* and *filter* methods was performed before the implementation of a Super-Learner for predicting past exposure to malaria vivax^13^. Here, the Least Absolute Shrinkage and Selection Operator (LASSO) regression was initially used to select one third of the responses. Then, using variable importance measures from Random Forest regression they iteratively selected the best responses which were then ranked by the *p-values* for the underlying Spearman’s correlation coefficient^13^. Although not implemented here due to the relatively small number of features, we envision that embedded feature selection approaches will be more useful in datasets in where the number of antibody responses exceeds the number of observations, as already seen in a study from Mali^14^. A forthcoming research study will investigate this solution and its impact on variable selection.

Alternative approaches to feature selection techniques for identifying the optimal antibody combinations for the task at hand have also been proposed^10,12^. These rely on simulated annealing algorithms that efficiently explore the vast space of feature combinations and thus identify the optimal feature combination solution given a fixed number of features defined by the user^10^. Whether this approach is preferable over feature selection techniques is an interesting research question for future work.

Concerning our predictive analysis, here we opted for the use of a SL^31^. The reasoning for this option relied on the fact that by combining the individual predictions of each classifier, the SL avoids the bias created by manually choosing the best-fitting model procedure and often provides better results than each individual classifier^31,32^. However, this was not always the case, as the Random Forest classifier alone tended to provide better predictions than the Super-Learner. Given that Random Forest is an *embeeded* method it performs feature selection during the classifier training procedure and thus we speculated that the removal of further features could be behind this increased performance^20,59^. Nevertheless, our validation analysis revealed that regardless of the strategy chosen for feature selection, nearly all features were important for classification purposes. This highlights the *filter* strategy’s ability to identify the most relevant features, avoiding any additional feature removal by the models embedded in the SL classifier.

However, this issue should be addressed in cases where the super SL embedded methods are implemented after a feature selection phase, such as done in Helb et al^13^, as further feature removal might occur without the user’s knowledge which may affect the interpretability of the results. Hence the slight decrease in the SL performance is expected to be explained by the SL attempt to correct for a possible overfitting to the data when using the Random Forest model. In this sense, these results should raise awareness concerning analysis where only the Random Forest approach is considered for predictive purposes, as it may lead to overfitting. Thus, the implementation of techniques such as the SL may provide more consensual results across the classifiers chosen for the predictive stage.

Comparing our results with the previous ones by Valletta and Recker^15^ revealed an increase in the prediction ability of up to 14% in the best-case scenario. This result further emphasizes the impact of feature selection prior to predictive analysis. On the one hand, this step removes antibody responses with negligible effect on clinical malaria. On the other hand, this stage decreases the number of features allowing for a more thorough feature analysis increasing the chance of finding the right transformation and dichotomization for each antibody response. Concerning the antibodies identified, we found that the antibody responses against different Merozoite Surface Proteins (MSPs) were consistently selected across the different feature selection strategies. These proteins are expressed at the parasite surface, thus, providing promising targets for malaria immunity because they are repeatedly and directly exposed to the host humoral immune system^7,8^. In particular, *msp2* has been associated with protection from clinical malaria in many studies and even suggested as a vaccine candidate^9–12^. For example, *msp2* has been strongly associated with protection against clinical malaria in two independent cohorts of Kenyan children^13^*. Msp4* has also been reported to have a protective effect in Kenyan children^14,16^. High antibody levels against *msp4* constructs have been associated with reduced morbidity in a Senegalese community^17^. *Msp7* protection against malaria has also already been identified in the Kenyan population^16,18^. Moreover, panels of antibodies comprising *msp7* have been associated with clinical protection against malaria in Kilifi, a rural district along the Kenyan coast^14^. In the same article, high antibody levels against the Erythrocyte-binding antigen-175 (*eba175*) antigen were also associated with protection from clinical malaria in children^14^ .Moreover, *eba175* is associated with protection from symptomatic malaria, as demonstrated in Papua New Guinean children^15^. These findings corroborate the ability of our methodologies to identify relevant antibodies associated with protection to malaria. However, *msp10* and *h103* have not been associated with clinical malaria protection. This evidence suggests that there are antibodies associated with protection against clinical malaria that have not yet been identified. Nevertheless, further studies are necessary to validate our findings. Finally, none of our feature selection metrics selected *msp1*, an immune response commonly associated with malaria protection and often referred to as potential vaccine candidates. Similar findings have been reported in other studies, where *msp1* has been described to show low or no associations with exposition or protection to clinical malaria^13,15^. These inconsistent findings further suggest the need for constructing robust feature selection strategies that could help increase reproducibility among studies. Nevertheless, one should consider the appropriateness of these feature selection strategies to identify antibodies associated with protection against clinical malaria, given that they were able to identify antibodies that have been constantly proposed for disease diagnosis.

## Conclusions

Here we have implemented feature selection strategies to analyze multiple antibody data. These were developed with the idea to couple classical, traditional statistical techniques for variable selection with popular machine learning techniques for predictive analysis. Considering the transformation of each antibody data individually these strategies represent a more flexible approach to accommodate different data patterns than the one commonly described in the literature. Overall, these methodologies led to an improved classification over previous analysis based on the use of the Random Forests alone, highlighting their potential to integrate future multi-sera pipelines.

### Abbreviation List

AIC: Akaike’s Information Criterion
Ama: Apical membrane antigen 1
AUC: Area Under de Curve
EBA: Erythrocyte-binding antigen
ELISA: Enzyme-linked immunosorbent assay
FDR: False discovery rate
GOF: Goodness of fitness
igG: Immunoglobulin G
LASSO: Least Absolute Shrinkage and Selection Operator
LDA: linear discriminant analysis
Log: logarithmic
LRM: Logistic regression model
MSP: Merozoite Surface Protein
MSRP: MSP7-related proteins
np: Number of Protected individuals
ns: Number of Susceptible individuals
Pf: Plasmodium *falciparum*
Prt: Protected
QDA: Quadratic discriminant analysis
RF: Random Forest
ROC: Receiver Operating Characteristic
rS: Spearman Correlation Coefficient
SeroTAT: serological testing and treatment
SL: super learner
sPLS-DA: Sparse partial least squares discriminant analysis
Sus: Susceptible
SVM: Support vector machine
SW: Shapiro-Wilk
χ^2^: Chi-square
XGB: Extreme Gradient Boosting

## Declarations

### Ethics approval and consent to participate

This study is based on publicly available data.

### Consent for publication

Not applicable

## Availability of data and materials

The datasets used and/or analyzed during the current study are available in this published article: Valletta, J. J. & Recker, M. Identification of immune signatures predictive of clinical protection from malaria. PLoS Comput Biol 13, e1005812 (2017). https://doi.org/10.1371/journal.pcbi.1005812. The R scripts generated are freely available in the following GitHub address: Immune-Stats (https://github.com/Publications/Fonseca_etal.).

## Authors’ contributions

AF and NS designed the work; AF and MS conducted the analysis; AF and NS interpreted the data; AF, PB, CC and NS have drafted the work or substantively revised it. AF, MS, PB, CC and NS have approved the submitted version.

## Data Availability

All data produced are available online at:https://doi.org/10.1371/journal.pcbi.1005812

## Acknowledgements

Not applicable

